# Determining the Optimal Sequence of Multiple Tests

**DOI:** 10.1101/2025.03.12.25323851

**Authors:** Lucas Böttcher, Stefan Felder

## Abstract

The use of multiple tests can improve medical decision making. The patient utility maximizing combination of these tests involves balancing the benefits of correctly treating ill patients and avoiding unnecessary treatment for healthy individuals against the potential harms of missed diagnoses or inappropriate treatments. We quantify the incremental net benefit (INB) of single and multiple tests by accounting for a patient’s pre-test probability of disease and the associated benefits and harms of treatment. We decompose the INB into two components: one that captures the value of information provided by the test, independent of the cost and possible harm of testing, and another that accounts for test costs and harm. We examine conjunctive, disjunctive, and majority aggregation functions, demonstrating their application through examples in prostate cancer, colorectal cancer, and stable coronary artery disease diagnostics. Using empirical test and cost data, we identify decision boundaries to determine when conjunctive, disjunctive, majority, or even single tests are optimal, based on a patient’s pre-test probability of disease and the cost-benefit tradeoff of treatment. In all three cases, we find that the optimal choice of combined tests depends on both the cost-benefit tradeoff of treatment and the probability of disease. An online tool that visualizes the INB for combined tests is available at https://optimal-testing.streamlit.app/.

## Introduction

Aggregating results from diagnostic and screening tests helps to improve overall test performance. ^5,8–10,15,16,22,25,28^ Different terms are used in the literature to describe various combinations of single tests. For instance, the protocol that classifies an individual as diseased if all tests return positive results is referred to as the “all heuristic” ^9,10^, “believe-the-negative rule” ^24^, “conjunctive positivity criterion” ^1,7,8^, and “orthogonal testing” ^14^. In Boolean algebra, this way of aggregating binary signals corresponds to using the binary AND operator. It implies that once a result is negative, testing stops and the patient remains untreated. Another aggregation method is referred to as the “any heuristic” ^9,10^ also known as the “believe-the-positive rule” ^24^ or the “disjunctive positivity criterion” ^1,7,8^. In this protocol, all tests must return negative results to classify an individual as healthy. Therefore, a single positive test is sufficient for a diagnosis, which in turn leads to treatment. In Boolean algebra, this aggregation method corresponds to the binary OR operator.

During the COVID-19 pandemic, various antigen and antibody tests were developed. ^6^ Similarly, multiple tests are available across various clinical settings, including diabetes testing ^4,11^, medical imaging ^3,32,35^, prostate cancer testing^29^, colorectal cancer testing^13^, and stable coronary artery disease testing ^17^.

With multiple tests available, how can one efficiently combine them to maximize their informational value? While efficient combinations are straightforward for two tests, calculations become increasingly complex as the number of tests increases. An algorithm has been proposed in the literature to combine test results and identify efficient combinations, using a knapsack-problem formulation.^10^ Another approach derives aggregated sensitivities and specificities from individual tests. ^2^ While both approaches can identify the receiver operating characteristic (ROC) frontier for combining constituent tests, neither provides a criterion for selecting the optimal combination.

The optimal test along a given ROC curve can be determined by considering the benefits of true positives, the utility loss from false positives, the cost of treatment, and the probability that the patient actually has the condition. ^7^ Interestingly, when the harm and cost of testing are taken into account, tests that are inefficient from an informational perspective (*i.e*., tests that fall inside the ROC curve) might still be optimal. Furthermore, the optimal combination of individual tests must also take into account their ranking order. Tests that are relatively cheap and harmless, and that lead to an early stop in testing due to the chosen positivity criterion may be prioritized.

To establish criteria for optimally aggregating test results, the remainder of this paper is organized as follows. The next section provides an overview of key parameters used to mathematically characterize the value of diagnostic information, as well as the benefits and risks associated with specific treatments. Subsequently, we derive the incremental net benefit (INB) for different test combinations and show how selecting the optimal test can be framed as a problem of maximizing this function. We then present three applications related to prostate cancer, colorectal cancer, and stable coronary artery disease diagnostics. For all three examples, we identify decision boundaries that determine when different combinations of tests should be used, depending on the cost-benefit tradeoff of treatment and a patient’s probability of disease. Finally, we discuss the findings and conclude the paper. An online tool that we developed to visualize the INB for various combinations of tests and parameters is available at https://optimal-testing.streamlit.app/.

## The Incremental Net Benefit of a Test

### The Treatment Threshold

We consider a diagnostic risk scenario in which uncertainty pertains to both a patient’s probability of illness and the potential benefits and harms of treatment. For an ill patient, a decision maker evaluates a treatment’s monetary net benefit as

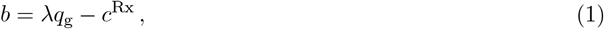

where *q*_g_ is the gain of quality-adjusted life years (QALYs), *λ* is the willingness to pay for a QALY, and *c*^Rx^ is the treatment cost. In contrast, a healthy patient will incur a monetary utility loss equal to

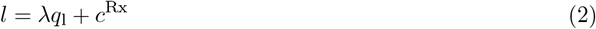

from the treatment. We now assume that the potential benefits, harms, and costs of treatment vary for each individual patient. A patient’s cost-benefit tradeoff associated with the treatment is represented by

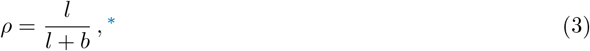

and the uncertainty about the health status is described by the pre-test probability of disease *p*, which also differs among patients. Facing a patient, characterized by (*p, ρ*), the decision maker evaluates the tradeoff between treatment and no treatment. The patient’s expected utility of treatment is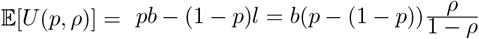, where we substituted 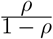 for *l/b* after the second equality sign. If this quantity is positive, treatment is recommended; otherwise, no treatment is preferable. This reasoning leads to the two treatment thresholds

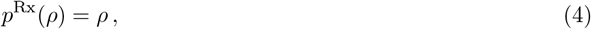

and

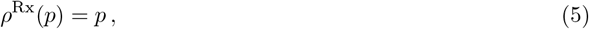

at which the decision maker is indifferent between treatment and no treatment ^†^ A patient should only be treated if *p* ≥ *p*^Rx^ or, equivalently, if *ρ* ≤ *ρ*^Rx^.

### The Value of Diagnostic Information

The treatment threshold *p*^Rx^ plays a central role in determining the informational value of a test, as it defines the decision maker’s choice in the absence of a diagnostic test. For a test with sensitivity Se (true positive rate) and specificity Sp (true negative rate), and a patient’s characteristics summarized by *p* and *ρ*, the value of diagnostic information is

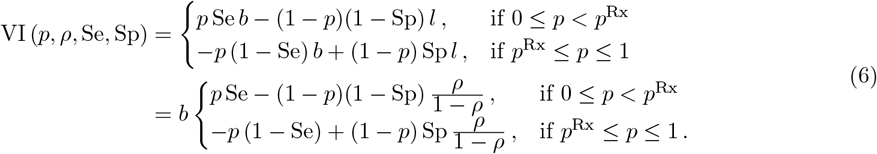

The function VI (*p, ρ*, Se, Sp) is the difference between the expected utility of the treatment decision with and without a test. Without a test, patients with a low probability of disease, *p*, would remain untreated. In contrast, with a test, patients with true-positive results receive treatment and gain utility, while those with false-positive results suffer a utility loss. In expected terms, the utility gain from true-positive outcomes is *p* Se *b*, while the utility loss from false-positive outcomes is (1 − *p*) (1 − Sp) *l*. Without a test, treatment is the preferred choice for patients with a high *p*. With a test, true-negative outcomes avoid the utility loss associated with unnecessary treatment, providing an expected benefit of (1 − *p*) Sp *l*. However, false-negative outcomes prevent the patient from receiving the benefits of treatment, resulting in an expected utility loss of −*p* (1 − Se) *b*.

### Test Thresholds

A test will come with monetary cost, *c*^Dx^, and potentially involve harm to the patient in case of invasive testing, represented by *λh*^Dx^. This leads to the concept of incremental net benefit (INB) of testing, defined as

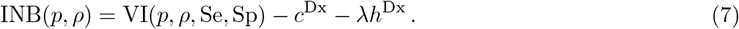

Using the INB, one can formulate the necessary and sufficient conditions for selecting the optimal diagnostic test. Let ℐ = {1, 2, …, *n*} be the set of all tests available for detecting a specific illness. The necessary condition for using test *i* ∈ ℐ is INB_*i*_(*p, ρ*) ≥ 0. The sufficient condition requires INB_*i*_(*p, ρ*) ≥ INB_*j*_(*p, ρ*) for all *i ≠j* ∈ℐ.

The literature on medical decision-making distinguishes between two approaches to defining the testing range. Pauker and Kassirer (1980) introduced the concept of a test interval for *p*, given *ρ*. ^21^ In contrast, Vickers and Elkin (2006) developed the decision curve analysis to determine the upper and lower bounds for *ρ*, given *p*. ^30^

By setting Eq. (7) equal to zero and solving for *p*, we obtain the corresponding test and the test-treatment thresholds

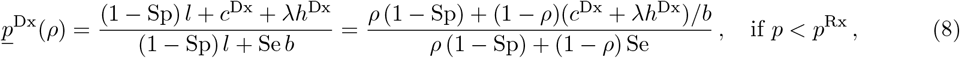

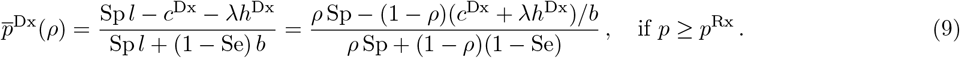

The test interval 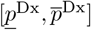 decreases if a test becomes more costly or more harmful. Starting from the INBs of two tests, *i* and *j ≠ i*, the probability of disease at which the decision maker shifts from preferring test *i* over test *j* is

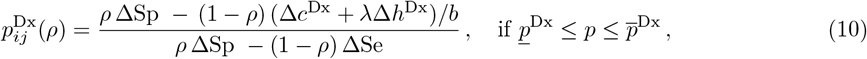

where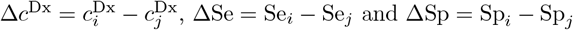.

In Figure 1, we show the INB of two tests as a function of the probability of disease *p*. The INB is linear in *p* and reaches its maximum value, *p J b* − *c*^Dx^, at *ρ*, where *J* = Se − (1 − Sp) is the Youden index. ^34^ The testing interval is determined by the minimum of the test thresholds and the maximum of the test-treatment thresholds. In the scenario shown in Figure 1, patients with 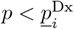 or 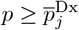 should not be tested. The former should not be treated and the latter undergo direct treatment. For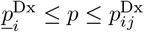, test *i* is recommended, and for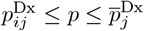, test *j* is preferred.

**Figure 1.**
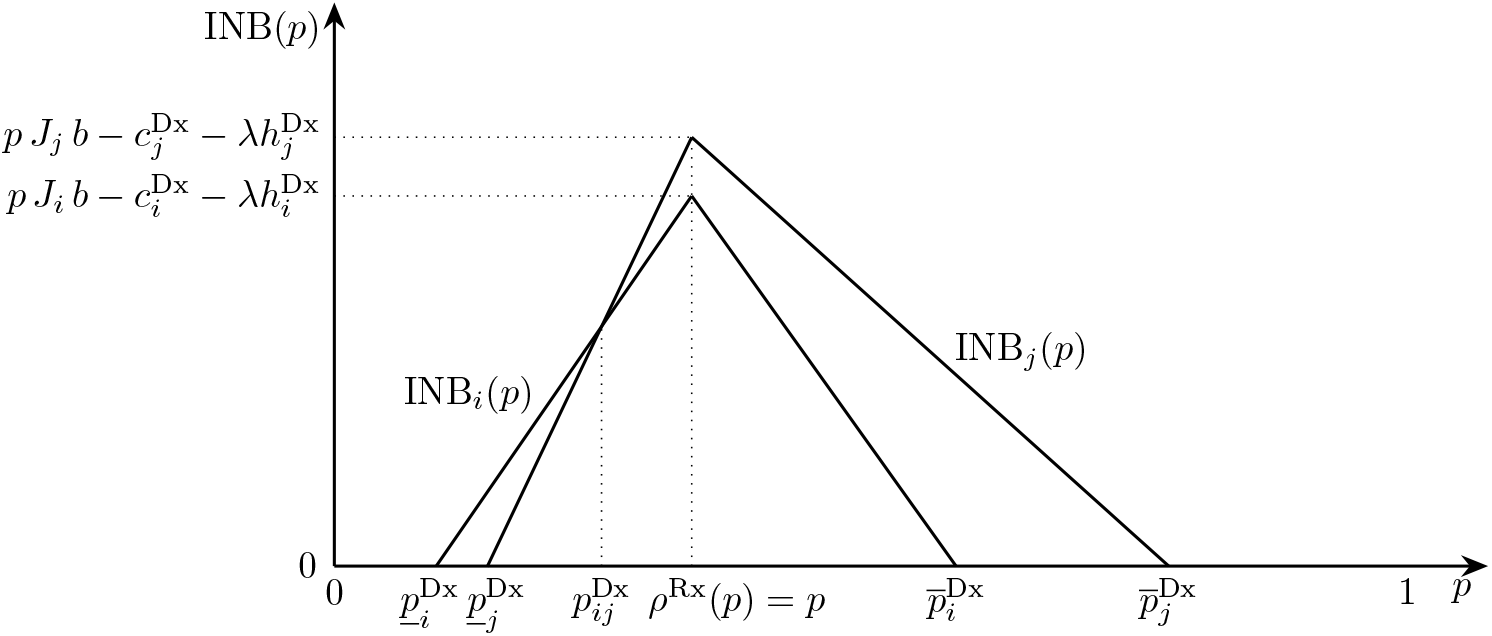
The incremental net benefits INB_*i*_(*p*) and INB_*j*_ (*p*) of two tests as a function of the probability of disease *p*, given *ρ*. The test and test-treatment thresholds are 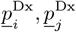 and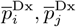, respectively. At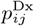, the decision maker shifts from preferring test *i* over test *j*.

Rather than expressing the thresholds in Eqs. (8)-(10) as a function of a patient’s cost-benefit tradeoff *ρ*, we can alternatively derive thresholds for *ρ*, given a patient’s probability of disease *p*. ^30^ Setting Eq. (7) equal to zero and solving for *ρ* yields a lower and upper bound

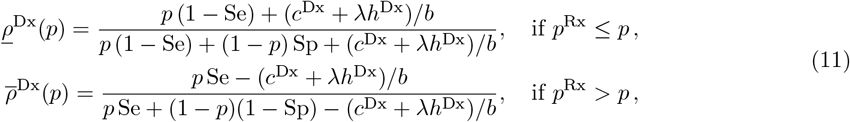

where a test with sensitivity Se, specificity Sp, cost *c*^Dx^, and harm *h*^Dx^ can be used.

The width of the test interval 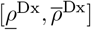 increases with *c*^Dx^ and *h*^Dx^. When the test is cost-free and causes no harm, the upper threshold corresponds to the positive predictive value, and the lower threshold corresponds to 1 minus the negative predictive value.

The treatment threshold at which the decision maker shifts from preferring test *i* over test *j* is

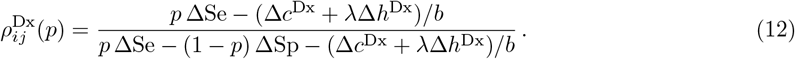

In Figure 2, we show the INB of two tests as a function of a patient’s cost-benefit tradeoff of treatment *ρ*. The INB is convex for *ρ < p* and concave for *ρ > p*. Given the probability of disease *p*, and the characteristics of the tests, including their costs, a testing range 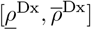 is defined. Patients for which 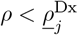 should be treated without prior testing, while for patients with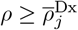, neither testing nor treatment is indicated. Patients in the range 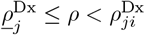 should undergo test *j*, and those in the range 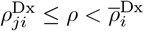 should receive test *i*.

**Figure 2.**
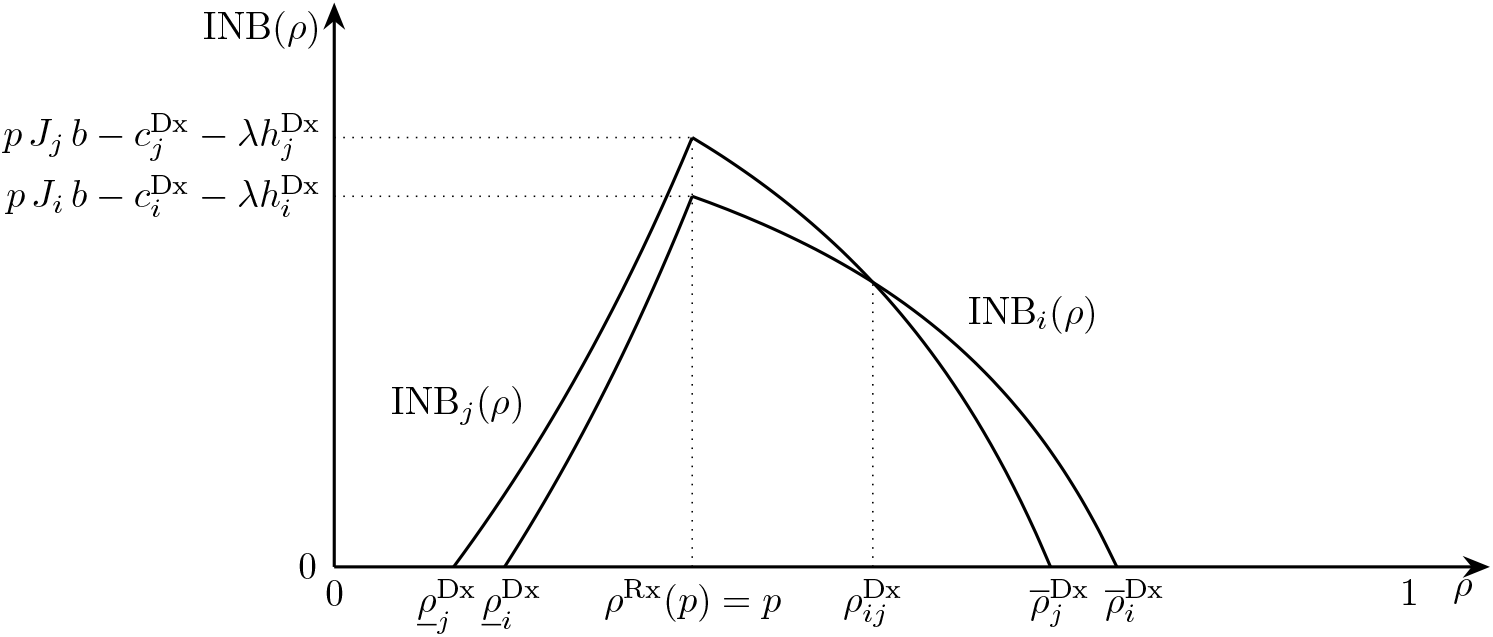
The incremental net benefits INB_*i*_(*ρ*) and INB_*j*_ (*ρ*) of two tests as a function of *ρ*. The lower and upper threshold were testing is indicated are 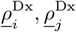 and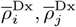, respectively. The probability of disease is *p*. At 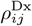, the decision maker shifts from preferring test *i* over test *j*.

### Sequencing Tests and Aggregating their Results

With multiple tests available, the decision maker must address the challenge of aggregating test results, choosing a positivity criterion, and determining the order in which the tests will be conducted. With a conjunctive positivity approach (using the AND operator in Boolean algebra), additional tests are applied if and only if the previous test yielded a positive result. In other words, the test sequence stops as soon as a negative outcome occurs. In contrast, with a disjunctive positivity approach (using the OR operator in Boolean algebra), further tests are performed if and only if the previous test is negative, meaning that testing stops as soon as a positive result is obtained.^‡^ With more than two tests available, a combination of the AND and OR operators and a majority criterion can be applied. The cost and harm of each individual test will play a key role in determining the specific sequence of tests.

In deriving our results, we assume that the outcomes of different tests are conditionally independent, given the disease status. This assumption is commonly used in the medical decision-making literature as it simplifies the mathematical analysis of aggregated test results. Additionally, manufacturers usually report performance measures for individual tests without specifying potential dependencies between them. However, in practice, test results may be correlated.

### Two Tests

The incremental net benefit of a sequence consisting of two tests, starting with test 1 and using the AND operator, is

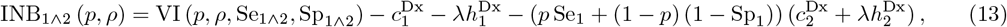

where 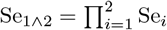 and 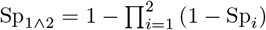. The sequence of tests (*i.e*., whether to start with test 1 or test 2) does not affect the value of diagnostic information; it only changes the expected testing cost. As *p* Se_1_ + (1 − *p*)(1 − Sp_1_) is the probability of a positive test outcome from test 1, test 1 has an advantage over test 2 not only if its cost and potential harm are lower, but also if it is expected to yield fewer (true and false) positive outcomes. This is because fewer positive outcomes make it less likely that test 2 will be needed.

The incremental net benefit associated with initiating the test sequence with test 1 and applying the OR operator is

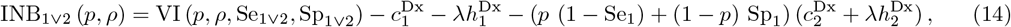

where 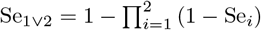 and 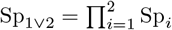. Again, the test sequence does not affect the value of diagnostic information. The expected cost and harm of test 2 depends on *p* (1 − Se_1_) + (1 − *p*) Sp_1_, the probability of a negative result from test 1.

### Three Tests

For three tests, the incremental net benefit associated with the AND operator is

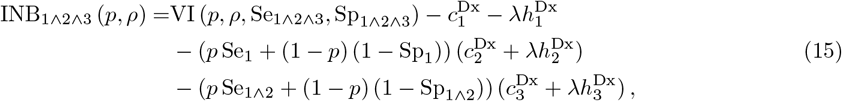

where 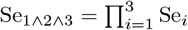 and 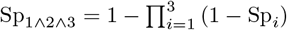.

Building on the INB from the two-test case [see Eq. (13)], we incorporate the term *p* Se_1∧2_ + (1 − *p*) (1 − Sp_1∧2_) = *p* Se_1_Se_2_ + (1 − *p*) (1 − Sp_1_) (1 − Sp_2_). This term accounts for the probability of a positive outcome after two tests, which leads to the use of the third test. As in the two-test examples, the specific test sequence does not affect the value of information; it only influences the expected cost and harm of testing.

For the OR operator, we have

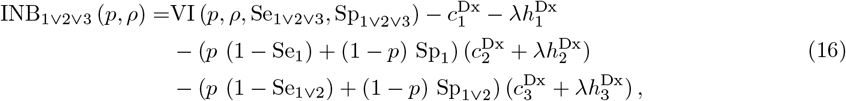

where 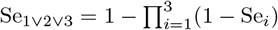 and 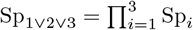. The probability of a negative outcome after two tests, which necessitates the use of a third test, is *p* (1 − Se_1∨2_) + (1 − *p*) Sp_1∨2_ = *p* (1 − Se_1_)(1 − Se_2_) + (1 − *p*) Sp_1_ Sp_2_).

With three tests, a test protocol based on the majority criterion offers another combinatorial option. If two tests yield positive outcomes, the decision maker would choose treatment, whereas two negative outcomes would lead to no treatment. The third test is required only when the first two tests produce conflicting results. This approach results in the incremental net benefit

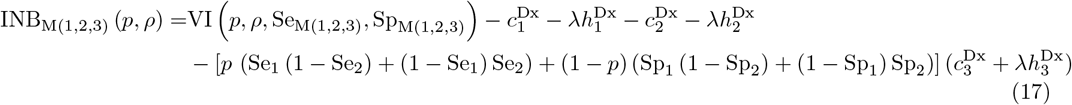

where

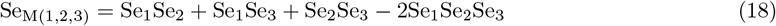

and

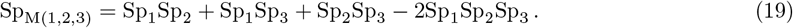

### AND and OR Aggregation of the Results of *n* Tests

Generalizing the previous equations to cases with *n >* 3 tests is straightforward. Adding another test affects the overall informational value of the test protocol, increases the expected cost including a potential harm in case of an invasive test, and may induce a further test, depending on the chosen positivity criterion. The incremental net benefit of a combined *n*-test, when using the AND operator, is

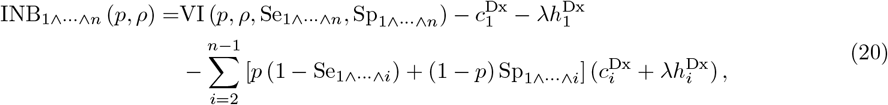

where

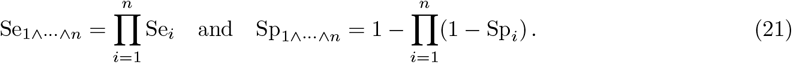

With the AND operator, overall sensitivity decreases, while overall specificity increases with *n*. In environments where the probability of disease is low, increasing the number of tests is appealing. A high specificity decreases the expected number of false positives, which is advantageous both from the informational and the cost perspectives. At the same time, applying one more test always implies an additional cost.

With the OR operator, we have

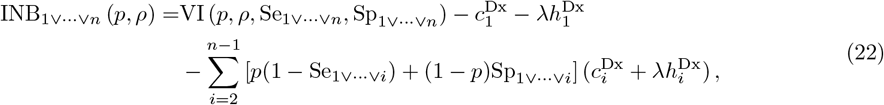

where

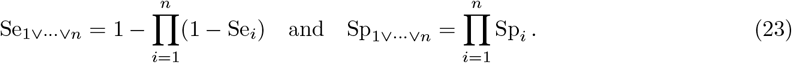

With the OR operator, overall sensitivity increases, while overall specificity decreases with *n*. If the probability of illness is high, decision makers will be inclined to increase the number of tests because a high sensitivity decreases the expected number of false negatives which is warranted both from the informational and the cost perspectives.

As shown in Eqs. (13)–(17), the value of diagnostic information does not depend on the order in which the *n* tests are conducted. Low-cost tests, when performed earlier in the sequence, are associated with a lower INB. Under the AND operator, tests that decrease the probability of positive outcomes are preferred due to their lower INB. In contrast, under the OR operator, tests that reduce the probability of negative outcomes are more likely to be prioritized earlier in the sequence.

For test strategies using the majority rule, we choose not to present the INB, overall sensitivity, or specificity when *n >* 3 and an odd number, as the corresponding mathematical expressions become very lengthy and their derivation is much more complex than for the AND and OR functions.

### Determining the Optimal Test

We use 𝒮 to denote a set of available single and combined tests. Given a patient’s pre-test probability of disease *p* and cost-benefit tradeoff *ρ*, the decision maker will select the test

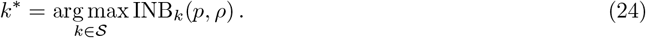

For the region in the (*p, ρ*) space where testing is indicated, the decision maker’s choice can also be described using the thresholds defined in the section “The Incremental Net Benefit of a Test”. For the set of available single and combined tests, we have

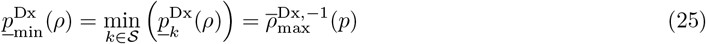

and

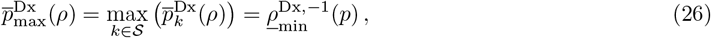

where 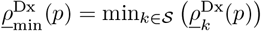 and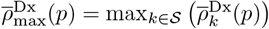. We use the notation 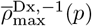 and 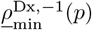 to indicate the inverse of 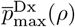 and 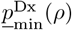, respectively.

This brings us to the following decision rules for a patient characterized by (*p, ρ*):

- Do not test or treat if 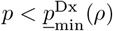 or, equivalently, 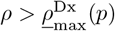
- Test if 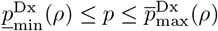 or, equivalently, 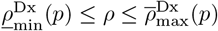
- Treat without testing if 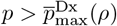 or, equivalently, 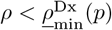

Within the region where testing is indicated, the optimal transition threshold can be determined by comparing all pairs 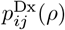 and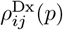. However, this approach requires quadratic memory and runtime, as every pair of tests must be evaluated, making it computationally complex. A more efficient method, linear in the number of tests, involves computing the envelope of INB_*k*_(*p, ρ*) and directly determining the optimal test and corresponding transition thresholds between tests using Eq. (24).

## Applications

We now turn to three applications to illustrate how the choice of the optimal test protocol varies with both the probability of disease *p* and the cost-benefit tradeoff of treatment *ρ*.

The first two examples focus on prostate cancer and colorectal cancer diagnostics, two diseases which exhibit a low prevalence. The third example considers stable coronary artery disease, a condition with relatively high prevalence in certain population groups. In all three cases, up to three tests can be combined using AND, OR, and majority functions.

### Prostate Cancer Diagnostics

Prostate-specific antigen (PSA) levels in the blood are used to identify men with prostate cancer. A cutoff of 20% for free-to-total PSA (FT) is applied to define a positive test result. Alternatively, or as a complement, human kallikrein 2 (hK2) can be used, with a cutoff set at 0.075 ng/mL. For these cutoffs, Vickers et al. (2013) report Se_FT_ = 0.91 and Sp_FT_ = 0.40 for FT, and Se_hK2_ = 0.51 and Sp_hK2_ = 0.78 for hK2. ^29^ Although the Youden index differs only slightly between the two tests (*J*_FT_ = 0.31 vs. *J*_hK2_ = 0.29), FT clearly dominates HK in terms of the positive likelihood ratio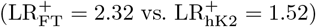. However, FT is inferior to HK with respect to the negative likelihood ratio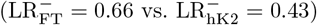 .^§^

A third option for prostate cancer testing is transrectal ultrasound (TRUS). For a cutoff of 50 cm^3^, Vickers et al. (2013) report Se_TRUS_ = 0.84 and Sp_TRUS_ = 0.34. The single tests and combined tests with varying sequences and positivity criteria result in 33 different test protocols (see Table 1). Since the sequence of tests does not affect the resulting sensitivity and specificity, these protocols produce 12 distinct pairs of sensitivity and specificity. From an information-theoretic perspective, nine of these test protocols are efficient, as they are part of the ROC frontier [see Figure 3(a)].^¶^ The individual tests FT, hK2, and TRUS, as well as the combined tests hK2 ∧ TRUS, hK2 ∨ TRUS, and FT ∨ TRUS, are not efficient as they are all weakly dominated by combinations of neighboring tests.

**Table 1.** Sensitivity and specificity of single and combined tests, as well as their optimal intervals, for prostate cancer diagnosis. We assume that 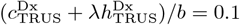 and 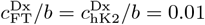

| Test | Se | Sp | LR<sup>+</sup> | LR<sup>-</sup> | Efficient test | Optimal test interval |  |
| --- | --- | --- | --- | --- | --- | --- | --- |
| | | | | | | $p^{\text{Dx}}(\rho = 0.26)$ | $\rho^{\text{Dx}}(p = 0.26)$ |
| <u>Single tests</u> |  |  |  |  |  |  |  |
| FT | 0.91 | 0.40 | 1.52 | 0.23 | no | [0.27, 0.44] | [0.12, 0.26] |
| hK2 | 0.51 | 0.78 | 2.32 | 0.63 | no |  |  |
| TRUS | 0.84 | 0.34 | 1.27 | 0.47 | no |  |  |
| <u>Combined tests</u> |  |  |  |  |  |  |  |
| AND ($n = 2$) | | | | | | | |
| FT, hK2 | 0.46 | 0.87 | 3.54 | 0.62 | yes | [0.12, 0.27] | [0.26, 0.53] |
| hK2, FT | 0.46 | 0.87 | 3.54 | 0.62 | yes |  |  |
| FT, TRUS | 0.76 | 0.60 | 1.90 | 0.40 | yes |  |  |
| TRUS, FT | 0.76 | 0.60 | 1.90 | 0.40 | yes |  |  |
| hK2, TRUS | 0.43 | 0.85 | 2.87 | 0.67 | no |  |  |
| TRUS, hK2 | 0.43 | 0.85 | 2.87 | 0.67 | no |  |  |
| AND ($n = 3$) | | | | | | | |
| FT, hK2, TRUS; FT, TRUS, hK2; hK2, FT, TRUS; hK2, TRUS, FT; TRUS, FT, hK2; TRUS, hK2, FT | 0.39 | 0.91 | 4.33 | 0.67 | yes |  |  |
| OR ($n = 2$) | | | | | | | |
| FT, hK2 | 0.96 | 0.31 | 1.39 | 0.13 | yes | [0.44, 0.63] | [0.1, 0.12] |
| hK2, FT | 0.96 | 0.31 | 1.39 | 0.13 | yes |  |  |
| FT, TRUS | 0.99 | 0.14 | 1.15 | 0.07 | no |  |  |
| TRUS, FT | 0.99 | 0.14 | 1.15 | 0.07 | no |  |  |
| hK2, TRUS | 0.92 | 0.27 | 1.26 | 0.30 | no |  |  |
| TRUS, hK2 | 0.92 | 0.27 | 1.26 | 0.30 | no |  |  |
| OR ($n = 3$) | | | | | | | |
| FT, hK2, TRUS; FT, TRUS, hK2; hK2, FT, TRUS; hK2, TRUS, FT; TRUS, FT, hK2; TRUS, hK2, FT | 0.99 | 0.11 | 1.11 | 0.09 | yes |  |  |
| Majority ($n = 3$) | | | | | | | |
| FT, hK2, TRUS; FT, TRUS, hK2; hK2, FT, TRUS; hK2, TRUS, FT; TRUS, FT, hK2; TRUS, hK2, FT | 0.88 | 0.55 | 1.96 | 0.22 | yes |  |  |

**Figure 3.**
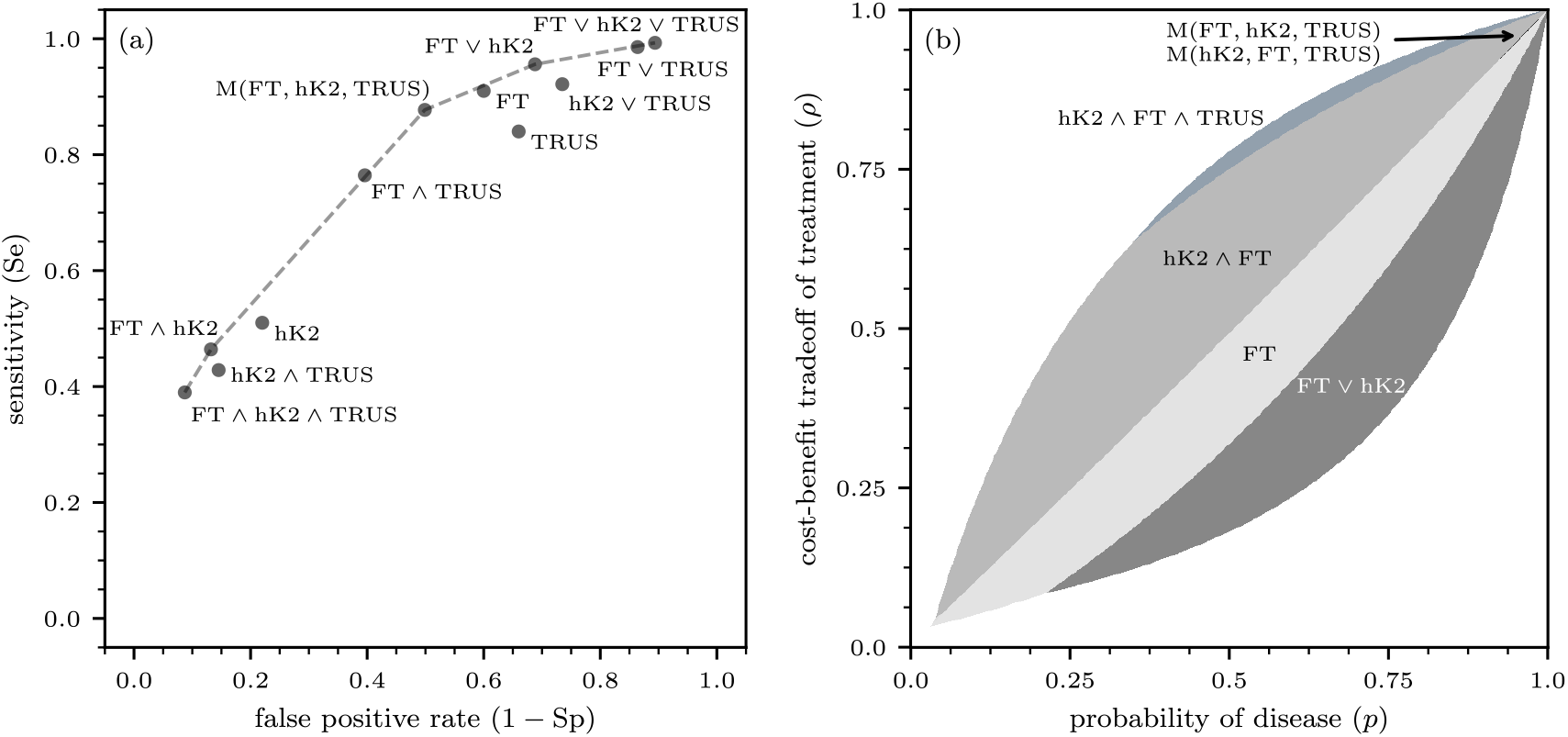
Prostate cancer diagnostics. (a) The ROC curve for prostate cancer testing. (b) Regions within the (*p, ρ*) unit square where different testing protocols are optimal. The sequence of tests does not influence the sensitivity and specificity values shown in the ROC plot. However, it is crucial in determining the optimal testing protocols illustrated in panel (b). Because the sequence of the first two tests in the majority protocol does not affect the outcome, the protocols M(FT, hK2, TRUS) and M(hK2, FT, TRUS) are equivalent in terms of their incremental net benefits.

TRUS involves inserting a probe into a patient’s rectum, which is uncomfortable for the patient and time-consuming for the physician. Vickers et al. (2013) quote an urologist who stated that he would perform no more than 10 ultrasound tests to detect cancer if the ultrasound was a perfect test. Assuming that this urologist anticipated the benefits, harm, and cost of a biopsy, as well as of the cancer treatment for true positives, we set *b* = 10(*c*^Dx^ + *λh*^Dx^). For their study on biopsy outcomes, Vickers et al. (2013) report that 26% of patients were positive for cancer.

For *ρ*^Rx^ = *p*, the informational value of a test is maximized. For *ρ*^Rx^ = 0.26, corresponding to a benefit-cost ratio of 2.85 in treatment, the overall test range for the pre-test probability of disease is [0.12, 0.63] (Table 1). The first optimal test within this range is the combined test hK2 ∧ FT, with 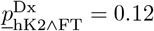. Among all single and double tests, it has the highest positive likelihood ratio. In the combined test hK2 ∧ FT, hK2 is performed first because its higher specificity compared to FT reduces the probability of positive test outcomes, and, consequently, decreases the probability that FT will be conducted. At 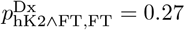 (above the treatment threshold), the single FT test begins to offer a greater incremental net benefit than hK2 ∧ FT. Notice that the single FT test has a very low negative likelihood ratio. At 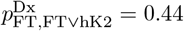, the combined test FT ∨ hK2, which has the lowest negative likelihood ratio, becomes the optimal testing protocol. The first test in this sequence is FT, which, due to its high sensitivity, reduces the probability of both negative test outcomes and the need for the second test. The testing range ends at 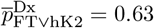.

With the probability of disease fixed at *p* = 0.26 and the cost-benefit tradeoff *ρ* varying, the range where testing is indicated is [0.11, 0.53]. The lower bound is reached by FT ∨ hK2, and the upper bound by hK2 ∧ FT. This corresponds to an interval of [8.09, 0.89] for *b/l* where testing is justified. At 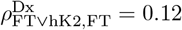, the single test FT becomes optimal. Then, at 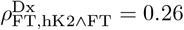 and for higher values of *ρ*, the conjunctively combined test hK2 ∧ FT is indicated. The corresponding benefit-cost ratio for FT and hK2 ∧ FT is 7.33 and 2.85, respectively.

Figure 3(b) shows the different test regions in the (*p, ρ*) space. Interestingly, for *p >* 0.3 and *ρ >* 0.65, the conjunctive triple test hK2 ∧ FT ∧ TRUS can be the optimal choice. However, the range of (*p, ρ*) combinations, where this is the case, is very narrow. The majority rule becomes a viable option only when *p* and *ρ* are around 0.9, where the benefit-cost ratio of treatment is 9.

Vickers et al. (2013) emphasize the importance of assessing the patient’s treatment preferences, which may be determined through a shared decision-making process. ^29^ They suggest that the typical *ρ* for prostate cancer biopsy is 20%, corresponding to a benefit-cost ratio of 4. As shown in Figure 3(b), this threshold roughly translates to a testing interval for *p* between 0.1 and 0.5.

Germany’s Robert Koch Institute (2022) reports the 10-year probabilities of developing prostate cancer for men at various ages: below 0.1% for those under 35 years, 0.4% at 45 years, 2.5% at 55 years, 6.2% at 65 years, and 6.7% at 75 years. ^27^ These probabilities are all below the minimum test threshold, indicating that men should not undergo single or combined tests for prostate cancer. Testing would only be reasonable for men over 65 if the benefit-cost ratio for biopsies did exceed 10. At this threshold, the combined test hK2 ∧ FT would be the preferred testing protocol due to its high positive likelihood ratio and low expected testing costs. A benefit-cost ratio of at least 24 for the biopsy followed cancer treatment would be required to justify using the single FT test alone.

### Colorectal Cancer Diagnostics

According to the Robert Koch Institute, the lifetime risk of developing colorectal cancer is approximately 1 in 25. Below age 65, the incidence rate is under 1%, but it increases to about 2% by age 80. ^26^ Many countries have implemented screening programs to detect colorectal cancer in the population. Several diagnostic options are available. The fecal immunochemical test (FIT) uses antibodies to specifically detect hemoglobin protein. Multitarget stool DNA testing (MTsDNA) identifies both hemoglobin and certain DNA biomarkers. Additionally, a colonoscopy examines the rectum, the sigma, and the entire colon using a flexible, lighted tube called a colonoscope. This device is equipped with a lens for viewing and a tool for tissue removal. While this invasive test is effective, it carries a perforation rate of 0.04% and, in the event of endoscopic perforation, a mortality rate of 7.5%. ^13^ With an assumed residual life expectancy of 15 years, the potential harm to the patient is equivalent to a loss of 0.0006 life years. Additional test characteristics, such as sensitivities and specificities, are summarized in Table 2, based on data from Pickhardt et al. (2003) and Ladabaum & Mannalithara (2016). ^13,23^

**Table 2.** Characteristics of FIT, MTsDNA, and colonoscopy for colorectal cancer diagnosis.

|  | FIT | MTsDNA | Colonoscopy | Treatment |
| --- | --- | --- | --- | --- |
| Se | 0.733 | 0.933 | 0.887 |  |
| Sp | 0.964 | 0.898 | 0.796 |  |
| $c^{Dx}$ | USD 19 | USD 649 | USD 1,400 | |
| $h^{Dx}$ | | | $6 \times 10^{-4}$ | |
| $c^{Rx}$ | | | | USD 75,000 |
| QALY |  |  |  | 1.5 |
| $b$ | | | | USD 75,000 |
| $c^{Dx}/b$ | $2.53 \times 10^{-4}$ | $8.65 \times 10^{-3}$ | | |
| $(c^{Dx} + \lambda h^{Dx})/b$ | | | $1.97 \times 10^{-2}$ | |

Figure 4(a) shows the ROC curve for colorectal cancer diagnostics. Similar to the previous example, for prostate cancer diagnostics, out of 33 combinations, 12 tests have distinct sensitivities and specificities. Five of these tests form the ROC frontier. None of the single tests belong to the frontier. Remarkably, the majority test protocol, with 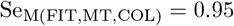 and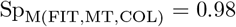, is very close to the maximum values of 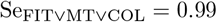 and 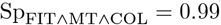.

**Figure 4.**
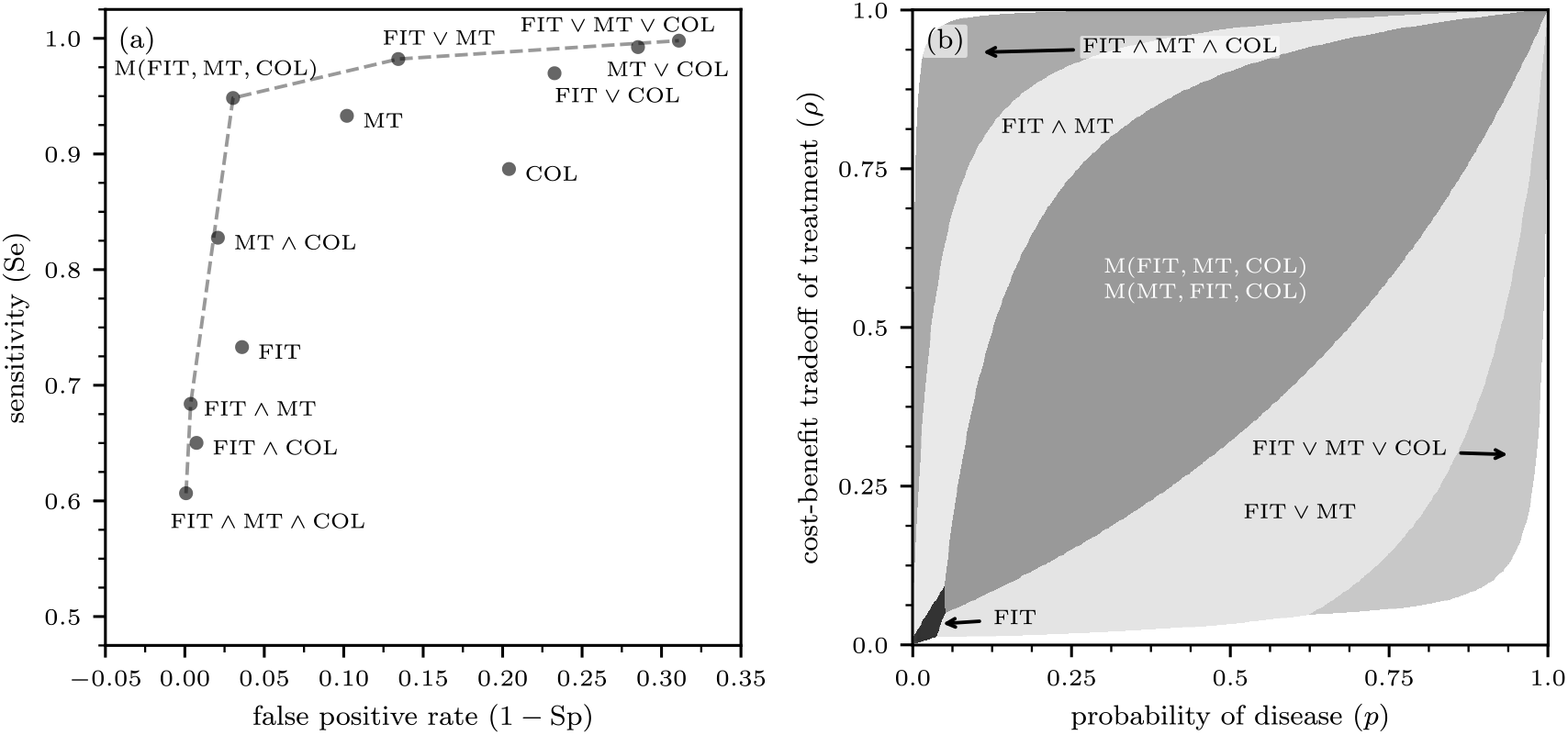
Colorectal cancer diagnostics. (a) The ROC curve for colorectal cancer testing. (b) Regions within the (*p, ρ*) unit square where different testing protocols are optimal. The sequence of tests does not influence the sensitivity and specificity values shown in the ROC plot. However, it is crucial in determining the optimal testing protocols illustrated in panel (b). Because the sequence of the first two tests in the majority protocol does not affect the outcome, the protocols M(FIT, MT, COL) and M(MT, FIT, COL) are equivalent in terms of their incremental net benefits.

Figure 4(b) shows the different test regions in the (*p, ρ*) space. Compared to the prostate cancer case, the overall area in which testing is indicated is significantly larger, primarily due to the higher accuracy of the tests for colorectal cancer. The single FIT test, which is not part of the ROC frontier, is the optimal test for low values of *p* and *ρ*. The combined test FIT ∨ MT is optimal for slightly larger values of *p*. For *p >* 0.05 and sufficiently large values of *ρ*, tests with majority aggregation function are optimal, provided that COL is used as the last test in the sequence. Whether to begin with FIT or MT makes no difference.

Given the low pre-test probability of colorectal cancer, only FIT and conjunctively combined tests with high specificities (Sp_FIT∧MT∧COL_ = 0.999 and Sp_FIT∧MT_ = 0.996) appear to be relevant in practice. For *p* = 0.02, the triple test is optimal if *ρ* ≥ 0.3, corresponding to a benefit-cost ratio for cancer treatment of 2.33. The side effects of colonoscopy are negligible, as the probability of requiring COL after a positive result for both FIT and MT is only 0.017. For a benefit-cost ratio between 2.33 and 20, the combined test FIT ∧ MT, with its higher sensitivity (0.68 vs. 0.61), becomes the optimal choice. For ratios exceeding 20, FIT alone is optimal, with a sensitivity of 0.73.

### Stable Coronary Artery Disease Diagnostics

The European Society of Cardiology (ESC) published guidelines on the management of stable coronary artery disease (CAD) in 2013. ^17^ These test guidelines differentiate according to a patient’s pre-test probability *p* of suffering stable CAD. The ESC task force recommended no testing if *p* is below 15%, and non-invasive testing in patients with *p* between 15% and 85%. If *p* exceeds 85%, the diagnosis of stable CAD should be made clinically.^?^ This recommendation is based on the observation that non-invasive cardiac tests on average have a sensitivity and a specificity equal to about 85%. The task force argues that because 15% of test results will be incorrect, not using a test at all will lead to fewer incorrect diagnoses for patients with *p <* 15% or *p >* 85%. Apparently, this recommendation does not consider the cost and potential harm of testing. Furthermore, it implicitly assumes that *b* = *l* (*i.e*., the net utility of treating a patient with stable CAD is equal to the utility loss of treating a patient without stable CAD).^∗∗^

In a recent publication, Min et al. (2017) analyzed single and combined test strategies for stable CAD, taking into account the cost and harm of testing and the benefit and cost of treatment. The different single tests include exercise treadmill testing (ETT), stress echocardiography (SE), myocardial perfusion scintigraphy (MPS), coronary computed tomographic angiography (CCTA), and invasive coronary angiography (ICA). ^18^ The latter, however, is rather costly and comes with a 1% mortality rate. Table 3 shows the parameter values based on data from Min et al. (2017). ^18^ MPS is dominated by SE and CCTA in terms of sensitivity, specificity, and cost. To calibrate the model, we set *b/l* = 1.9 such that the test threshold for ETT is equal to 15%. At the same time, the test-treatment threshold for CCTA becomes 87%, which is close to the ESC 2013 guidelines at which non-invasive testing is no longer indicated. The average cost of treatment, estimated by Min et al. (2017), is *l* = USD 55,000 for patients with a 20% pre-test probability of stable CAD.

**Table 3.** Characteristics of ETT, SE, MPS, and CCTA for stable CAD diagnosis.

|  | ETT | SE | MPS | CCTA | ICA and Rx |
| --- | --- | --- | --- | --- | --- |
| Se | 0.68 | 0.867 | 0.806 | 0.937 |  |
| Sp | 0.77 | 0.807 | 0.747 | 0.847 |  |
| $c^{\text{Dx}}$ | USD 100 | USD 340 | USD 819 | USD 394 | |
| $l$ | | | | | USD 55,000 |
| $b$ | | | | | USD 105,000 |
| $c^{\text{Dx}}/b$ | $9.57 \times 10^{-4}$ | $3.25 \times 10^{-3}$ | $7.84 \times 10^{-3}$ | $9.57 \times 10^{-4}$ | |

According to Figure 5(a), five different test strategies constitute the ROC curve for stable CAD testing. The single tests SE and ETT are far off the efficient frontier. CCTA is weakly dominated. The highest sensitivity is achieved with the disjunctive triple test ETT ∨ SE ∨ CCTA, the highest specificity with the conjunctive triple test ETT ∧ SE ∧ CCTA. The double tests that exclude the inefficient ETT, *i.e*., SE ∨ CCTA and SE ∧ CCTA, are also part of the ROC curve, as is the triple test with the majority rule.

**Figure 5.**
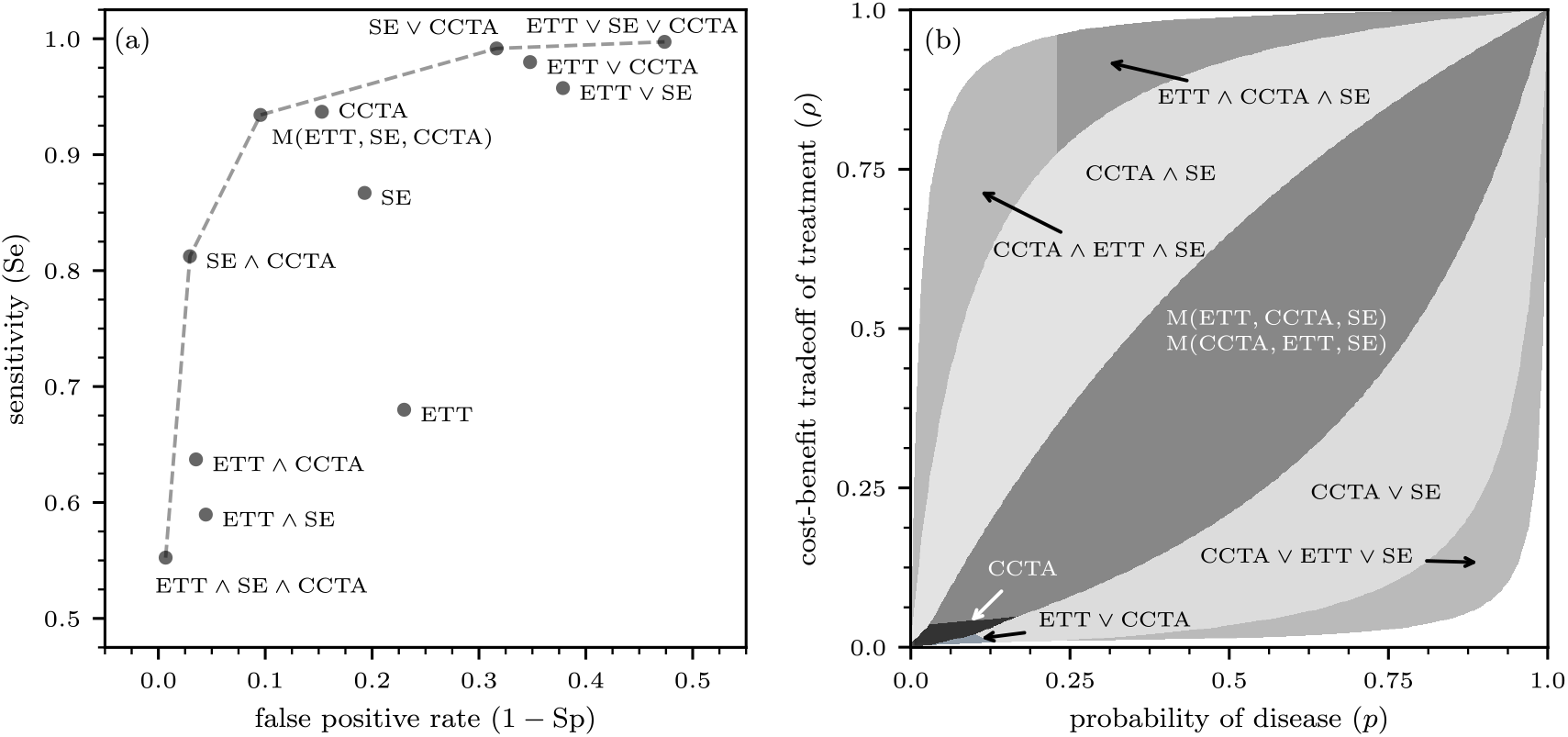
Stable coronary artery disease (CAD) diagnostics. (a) The ROC curve for CAD testing. (b) Regions within the (*p, ρ*) unit square where different testing protocols are optimal. The sequence of tests does not influence the sensitivity and specificity values shown in the ROC plot. However, it is crucial in determining the optimal testing protocols illustrated in panel (b). Because the sequence of the first two tests in the majority protocol does not affect the outcome, the protocols M(ETT, CCTA, SE) and M(CCTA, ETT, SE) are equivalent in terms of their incremental net benefits.

Figure 5(b) shows the optimal test protocols, depending on a patient’s probability of stable CAD, *p*, and their individual treatment threshold *ρ*. The single tests ETT and SE are never optimal. CCTA is the best option if both *p* and *ρ* are low, specifically when *p <* 18% and 1% *< ρ <* 3%. If *ρ <* 1%, the disjunctive double test ETT ∨ CCTA can be the preferred choice. CCTA would only be used if ETT is negative. Despite its insufficient test accuracy, ETT is used first because it is much less costly than CCTA.

The task force also published testing ranges for individual test options. If the patient is suitable and the technology as well as the local expertise is available, the ESC guidelines recommend the use of CCTA in patients at low to intermediate *p* of 15–50%. Alternatively, for patients with *p* between 15–85%, stress imaging testing (SE, MPS, SPECT, PET) is advised. If we follow the task force’s implicit *ρ* = 0.5, CCTA is optimal for *p* up to 50%, although not as a single test, but in conjunctive combination with SE. CCTA again is optimal for high *p*, now in disjunctive combination with SE. Changes in *ρ*, including down to 35%, which follows from *b/l* = 1.9, will not change the optimal test strategies as a function of *p* much. Given the relatively wide range of *p* for patients suffering from stable CAD, the majority functions M(ETT, CCTA, SE) and M(CCTA, ETT, SE) may be optimal, depending on the values of *p* and *ρ*. Because the sequence of the first two tests in the majority protocol does not affect the outcome, the protocols M(ETT, CCTA, SE) and M(CCTA, ETT, SE) are equivalent in terms of their incremental net benefits.

## Discussion

We studied the optimal aggregation of results from multiple diagnostic tests, using the incremental net benefit (INB) to quantify the tradeoffs between the informational value of the tests, test costs, and the associated benefits and harms of treatment. An online tool that visualizes the INB for various combined tests and parameters is available at https://optimal-testing.streamlit.app/.

Consistent with prior work on aggregating the results of multiple tests ^2,10^, our findings confirm that the receiver operating characteristic (ROC) curve is useful for evaluating tests based on their informational value. However, an efficient test (*i.e*., one located on the ROC frontier) may not be optimal for a specific medical application, as optimality requires maximizing the INB, which depends on both the test’s informational value and health-economic factors. Likewise, tests that are inefficient from an informational perspective may still be optimal due to their low costs and minimal side effects.

Using three application examples focused on prostate cancer, colorectal cancer, and stable coronary artery disease diagnostics, we identify decision boundaries that determine when different combinations of tests are optimal, based on a patient’s pre-test probability of disease and their cost-benefit tradeoff from treatment. For prostate cancer diagnostics, the most relevant tests are the free-to-total prostate-specific antigen (PSA) test and its combination with the human kallikrein 2 (hK2) marker, where hK2 is performed first, and the PSA test is conducted if the HK result is positive. However, the benefit-cost ratio of a biopsy in case of positive test outcomes needs to be 10 to justify the use of the combined double test and even 24 for the single hK2 test. The implied small range for testing for prostate cancer is due both to the low accuracy of these tests and the low prevalence of this cancer. For colorectal cancer, the single fecal immunochemical test and conjunctively combined triple tests are particularly relevant due to the disease’s low prevalence. In contrast, for stable coronary artery disease, a broader range of tests, including the single coronary computed tomographic angiography test, conjunctively and disjunctively combined triple tests, and majority protocols, is practically relevant due to the condition’s wider prevalence range.

Two limitations are worth noting. First, we assume that the outcomes of different tests are conditionally independent, given the disease status. This assumption is widely used in the medical decision-making literature as it simplifies the mathematical analysis of aggregated test results. Moreover, manufacturers typically report performance measures for individual tests without addressing potential dependencies between them. However, in practice, test results may exhibit correlations. Second, while we used established estimates for parameters such as test costs and the benefits and harms of treatment, these parameters may vary in practice due to heterogeneous population effects and other context-specific factors.

Both limitations present valuable opportunities for future research. Quantifying the effects of correlations between test results and obtaining more accurate estimates for the parameters involved in the INB calculation can contribute to further improving medical decision-making processes that rely on aggregating results from multiple tests. Another interesting direction for future work is to study the applicability of our proposed methods in infectious disease monitoring and management.^19,33^

## Data Availability

An online tool that visualizes the INB for combined tests is available at https://optimal-testing.streamlit.app/.

## Acknowledgments

The authors thank Luis L. Fonseca, Bernt-Peter Robra, Kevin Selby, Dilek Sevim, and Michael Zellweger for helpful comments. Financial support for this study was provided in part by a grant from hessian.AI and the Army Research Office (W911NF-23-1-0129) to LB.

## Footnotes

‡ In the following sections, we use the notations *x ∧ y* and *x ∨ y* to represent the Boolean operations *x* AND *y* and *x* OR *y*, respectively.

§ LR^+^ = Se*/*(1 *−* Sp), LR^−^ = (1 *−* Se)*/*Sp.

3 ^¶^If arbitrary AND/OR combinations are allowed, the tests (FT *∧* hK2) *∨* (FT *∧* TRUS) with Se = 0.88 and 1–Sp = 0.44, and FT *∨* (hK2 *∧* TRUS) with Se = 0.95 and 1–Sp = 0.66, are efficient. As a result, FT *∧* TRUS would become inefficient.

∥ ESC refined its guidelines in 2019 and 2024. ^12,31^ By and large, it confirmed the *>*15%–85% non-invasive testing range for the pre-test probability, although it narrowed the targeted indication from “stable CAD” to “obstructive CAD”.

∗∗ This can be verified if we set *p*^Dx^ = 0.15, *p*^Dx^ = 0.85 and solve for *b/l* [see Eqs. (8) and (9)].

